# Disruption of functional gradients in genetic generalized epilepsy and its clinical relevance: evidence from high-density EEG

**DOI:** 10.1101/2025.09.09.25335308

**Authors:** Juanli Zhang, Isotta Rigoni, Dimitri Van De Ville, Serge Vulliémoz, Nicolas Roehri

## Abstract

Genetic Generalized Epilepsy (GGE) represents around 20% of adult epilepsies and involves widespread network dysfunction across both hemispheres. While abnormalities across functional networks are well-documented, the hierarchical organization of these networks in GGE remains unknown. Our goal was to investigate whether GGE alters this hierarchy estimated using gradient analysis based on EEG-informed connectome. We analyzed a high-density EEG dataset from 20 GGE patients and matched healthy controls (HC). Compared to HC, GGE patients had significantly less negative gradient scores in the frontoparietal network (FPN) in the secondary gradient of the beta frequency band (p=0.0201, FDR corrected, quantified as large effect size by Cliff’s delta: 0.52). Additionally, in the theta band, the secondary gradient scores in multiple networks were closely associated with epilepsy duration (Spearman’s |*R*|>0.50, all p<0.05, FDR corrected). The findings were robust for different thresholds and not explained by potential confounders. In GGE, FPN moving closer to the other networks might promote the widespread pattern of pathological activity, and the association between gradient scores and epilepsy duration supports a progressive disruption of the network gradients. Altogether, this study presents the first EEG-based evidence of GGE-related gradient signatures and its clinical relevance.

**Author Summary:** GGE represents around 20% of adult epilepsies and involves abnormalities across multiple networks. However, the hierarchical organization of the networks, which could be captured by gradient analysis, associated with GGE remains elusive. Here, we derived the frequency-dependent gradient patterns from high-density EEG of GGE patients and matched healthy controls. In GGE patients, beta band secondary gradient (G2) showed significantly less negative gradient scores in the frontoparietal network compared to HC, indicating a narrowing network hierarchy which might promote the widespread pattern of pathological activity. Additionally, theta band G2 of multiple networks were closely associated with epilepsy duration, suggesting a progressive disruption of network hierarchy as epilepsy advances. This study presents the first EEG-based evidence of GGE-related gradient signatures and its clinical relevance.

## Introduction

While focal epilepsy is more common, generalized epilepsy, where the seizure onset involves both hemispheres and multiple brain regions, makes up around 30% of the cases^1^. Most generalized epilepsies, classified as Genetic Generalized Epilepsy (GGE), have a presumed genetic etiology and feature polygenetic inheritance^2^. In terms of electrophysiology, GGE shows widespread cortical activity including the generalized (poly)spike-and-wave (GSW) discharges in the interictal states. Although interictal GSW is a key diagnostic marker^3^, it may be absent or rare in some cases, highlighting the need for identification of additional EEG markers.

GGE is considered a fronto-cortico-thalamic circuit disorder^4,5^, involving abnormal engagement of multiple regions and networks. Previous studies show that patients with GGE exhibited altered intrinsic activity across higher-order (e.g., default mode network (DMN) and attention network) and low-level perceptual systems (e.g., somatomotor and visual networks), as well as disrupted inter-network communication^6–8^. Although brain network’s topology in GGE remains insufficiently characterized at the global level^9,10^, our prior work based on the same high-density EEG (hdEEG) dataset suggests that regional re-organization may compensate to preserve global topology^11^. Altogether, these findings highlight distributed abnormalities and functional re-organization in GGE. Meanwhile, parallel works propose that the brain functional organization follows smooth spatial gradients where nearby regions connect to nearby areas so that they exhibit graded, overlapping connectivity patterns (heterogeneity and multiplicity)^12–14^. However, whether GGE disrupts this orderly spatial arrangement remains unknown.

One appealing approach to fill this gap is projecting the high-dimensional functional connectome onto low-dimensional representations termed gradients. This data-driven approach maps the connectivity patterns onto low-dimensional manifolds capturing the continuum from unimodal to transmodal regions and revealing shifts in network hierarchy^15–18^. Regions with similar connectivity profiles are located at similar positions along the gradient axes, while those with dissimilar connectivity profiles are positioned further apart. For fMRI, gradient representations have been well established^16, 19–21^; however, EEG-informed gradients have not yet been reported. Our study therefore aims to: 1) derive EEG-derived functional gradients, and 2) characterize the network re-organization along these axes in GGE patients.

Using hdEEG-derived functional connectomes, we investigated frequency-specific gradient alterations in GGE patients by examining: 1) functional network reorganization through gradient patterns, and 2) clinical correlations between gradient scores with epilepsy progression. Additionally, we verified the robustness of results via threshold-varied analysis and confound-control analyses.

## 2 Materials and Methods

### 2.1 Participants

The dataset used in this study was originally collected and preprocessed for a previous investigation^11^. Briefly, a group of patients diagnosed with GGE (n=20, 12 females, median age=31.5 years, age range of 18-57 years, median epilepsy duration of 15.5 years) and a group of sex- and age-matched (10 females, median age=31 years, age range of 23-54 years) healthy controls were included retrospectively and prospectively through the database of the Geneva University Hospital for this study. All the included participants were over 18, without prior neurosurgery, multifocal epileptic, or focal MRI (magnetic resonance imaging) abnormality.

### 2.2 EEG acquisition and pre-processing

Participants underwent hdEEG recording at 1000 Hz (256 electrodes, Electrical Geodesic System) while calm and awake with eyes closed. The patients were recorded for ∼20 minutes whilst the healthy controls (HC) for 10 minutes to ensure sufficient data after removing the segments with interictal epileptiform discharges (IEDs, marked by an expert epileptologist).

Data were first down-sampled to 250 Hz, and band-pass filtered (1-45 Hz). An additional notch filter (50 Hz) was applied and independent component analysis was performed to remove the artifactual sources of oculomotor, cardiac, and muscle activity. Further, bad channels were interpolated with neighboring ones and the data were re-referenced to the common average. All the pre-processing steps were implemented in the Fieldtrip Toolbox^22^ in Matlab and are more extensively described in the previous study^11^.

### 2.3 MRI imaging and source reconstruction

Individual structural T1-weighted MRI (3T, Siemens Prisma) was utilized for individual head modeling and source imaging. EEG forward model employed a three-layer BEM approach with the default conductivity settings using OpenMEEG^23^. Next, approximately 5000 source points without constrained orientation were distributed within the grey matter. Inverse solution was computed with eLORETA (exact low resolution brain electromagnetic tomography)^24^.

The source space was parceled into 460 regions of interest (ROI) according to the 4th scale of the Lausanne atlas (version 2018)^25^. Note, during the source imaging, sub-cortical structures of both hemispheres, namely thalamus, pallidum, caudate, nucleus accumbens and putamen, were already excluded due to the uncertainty in source imaging precision, retaining in total 450 ROIs. Within each ROI, ROI time series was derived via singular value decomposition by decomposing and projecting data based on the principal component^26^.

### 2.4 Functional connectivity

Functional connectivity between ROI was quantified using the debiased weighted phase lag index (wPLI)^27^ for its robustness to source leakage when measuring the phase synchrony through weighting the observations by the magnitude of imaginary component of the cross-spectrum. wPLI was computed between 1-40 Hz (0.5 Hz steps) using a 2s sliding window with 50% overlap. A functional connectome was obtained for each frequency band by averaging the connectivity values within the specific band: 1-4 Hz for delta, 4.5-7 Hz for theta, 7.5-12.5 Hz for alpha and 13-30 Hz for beta bands. Windows containing artifacts (segments exceeding the absolute threshold 120µV or corresponding to IED) were removed, and a maximum of 450 windows per participant were randomly selected to minimize the length bias. Given prior strong evidence for network re-organization effects in theta and beta bands^11^, we hypothesized that these frequency bands could be our prior interest. Nevertheless, other canonical frequencies were explored and briefly reported.

### 2.5 Gradient estimation for the ROI and networks

The estimation of gradients was performed following established approaches^14,16,28^. For each participant, we thresholded (row-wise) functional connectomes (450×450 matrix per frequency band) to retain the top 20% connections (using 80^th^ percentile). We computed an affinity matrix (representing the connectivity similarity profiles) using cosine similarity of the thresholded connectome, then applied diffusion mapping, a non-linear dimensionality reduction technique, to embed the high-dimensional data into a low-dimensional space while preserving intrinsic geometry. This non-linear approach identifies the principal axes (or gradients) explaining the largest variances via the eigen-decomposition of the graph Laplacian (which is not a feature of linear methods such as PCA) of the affinity matrix. The density parameter *α* was set to 0.5 for a balanced global structure preservation and noise robustness in the diffusion mapping, following established recommendations^16,29,30^. All steps were implemented using BrainSpace^18^.

To overcome the randomness of direction when yielding gradients, we created a group template from an “out-of-sample” dataset consisting of 137 healthy subjects (median age=32 years, 64-channel recording, no age difference (p=0.13))^31,32^ by averaging the functional connectivity matrices extracted as described above. Individual gradients from this study were aligned to the group template via Procrustes rotation^16,18^. The first two gradients, the principal (G1) and secondary (G2) gradients, explaining around 16% and 10% of the variances, respectively, were analyzed and reported.

Network-level gradient scores were derived by averaging across the ROI values within each canonical Yeo7 functional network^33^.

While primary analyses used 80th percentile threshold, we tested robustness across a family of thresholds (30^th^-90^th^ percentiles in 10% step to retain top 70%-10% connections).

### 2.6 Statistical analysis

The gradients scores were compared for each of the seven networks between two groups (HC versus GGE) using permutation test to ensure the robustness against non-normality. For each permutation, the label of the subjects was randomly shuffled and the difference of the means from two groups was computed. In total, 2000 permutations generated a permutation distribution to which the actual differences of the means were compared to. To find a p-value, we simply computed the proportion of permutations where absolute values of the differences in the means exceeded absolute value of the observed difference in means.

False discovery rate (FDR) correction was applied for multiple comparisons (7 comparisons)^34^. All the tests were reported after the FDR-correction. A non-parametric effect size measure, Cliff’s delta, was applied to quantify the effect size.

To investigate the clinical relevance of the gradients in GGE patients, we calculated the correlation between the gradient scores with the epilepsy duration while controlling for age (partial correlation, type “Spearman”). Analogously, we also computed the correlation between gradient scores and drug load (sum of the ratios between actual dose and defined daily dose over all the antiseizure drugs)^35^. Spatial consistency across all the thresholds was evaluated via Spearman correlation of group-averaged gradient maps across ROIs.

## 3 Results

### 3.1 Template of gradient from EEG-based functional connectome

While well-established fMRI gradients maps exist, no such maps have been developed based on functional connectome from EEG. To this end, a set of gradients was derived from the group-level averaged functional connectome based on a large-sample open dataset^32^. As shown in Figure 1, the template gradients from the theta and beta frequency band showed some recognizable patterns. The principal theta gradient seemed to follow a posterior-to-anterior axis, whilst the secondary gradient aligned with a medial-to-lateral transition. Similarly, the beta band’s G1 ordered the nodes along the posterior-to-anterior direction. In contrast, the G2 seemed to align along a left-posterior to right-anterior-central axis.

**Figure 1.**
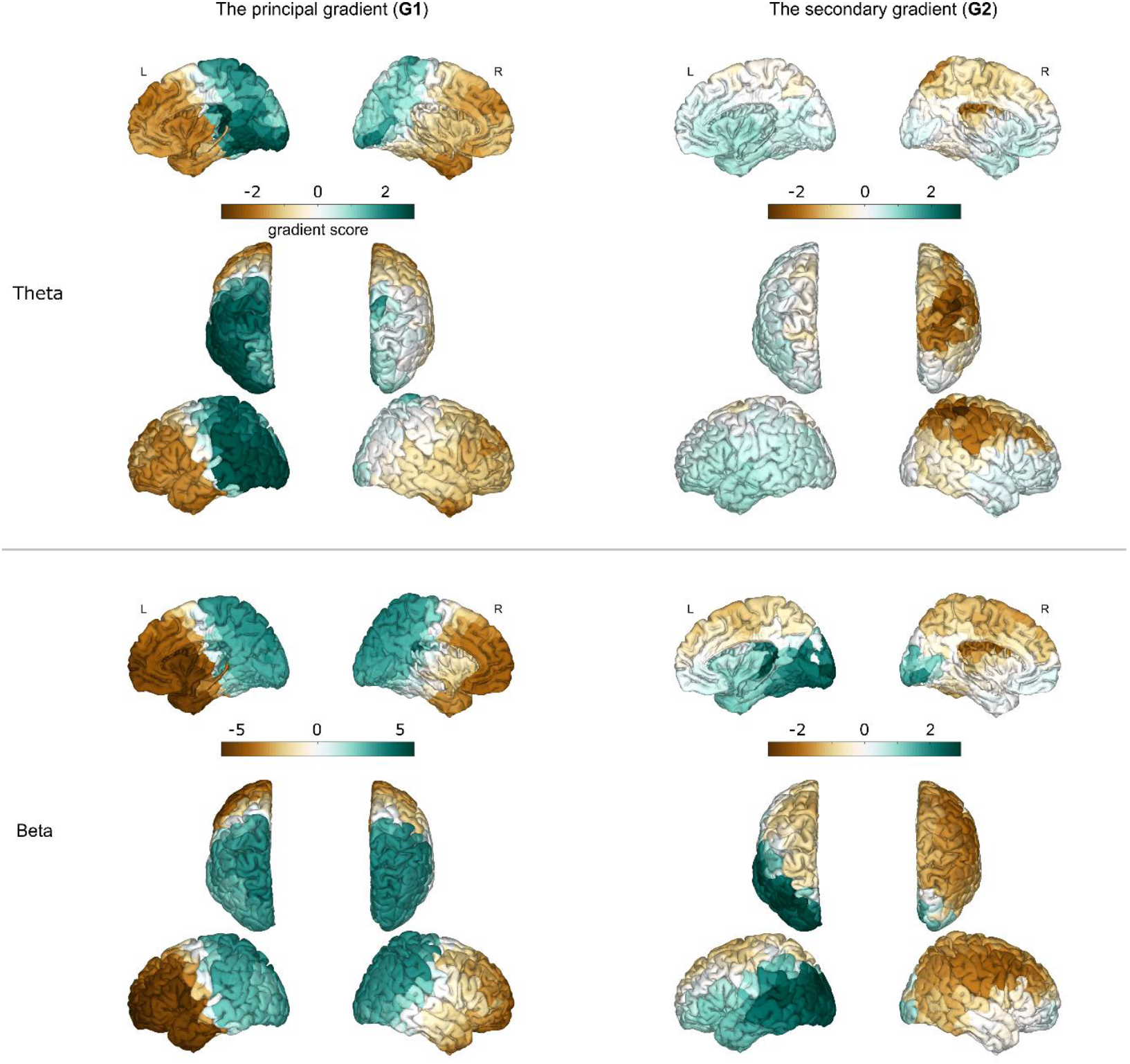
The template gradients from an out-of-sample open dataset. Topographies show the gradient distributions from theta (upper panel) and beta (lower panel) frequency bands for both the principal (G1) (left column) and the secondary (G2) components (right column). The color bar represents the gradient score.

### 3.2 Network-level principal and secondary gradients

After aligning each individual’s gradients to the group template, group-average gradients (G1 and G2) were obtained for display purposes by averaging the gradient scores across all the subjects within each group in the theta and beta bands. The gradient patterns (Figure 2) closely matched the template (Figure 1).

**Figure 2.**
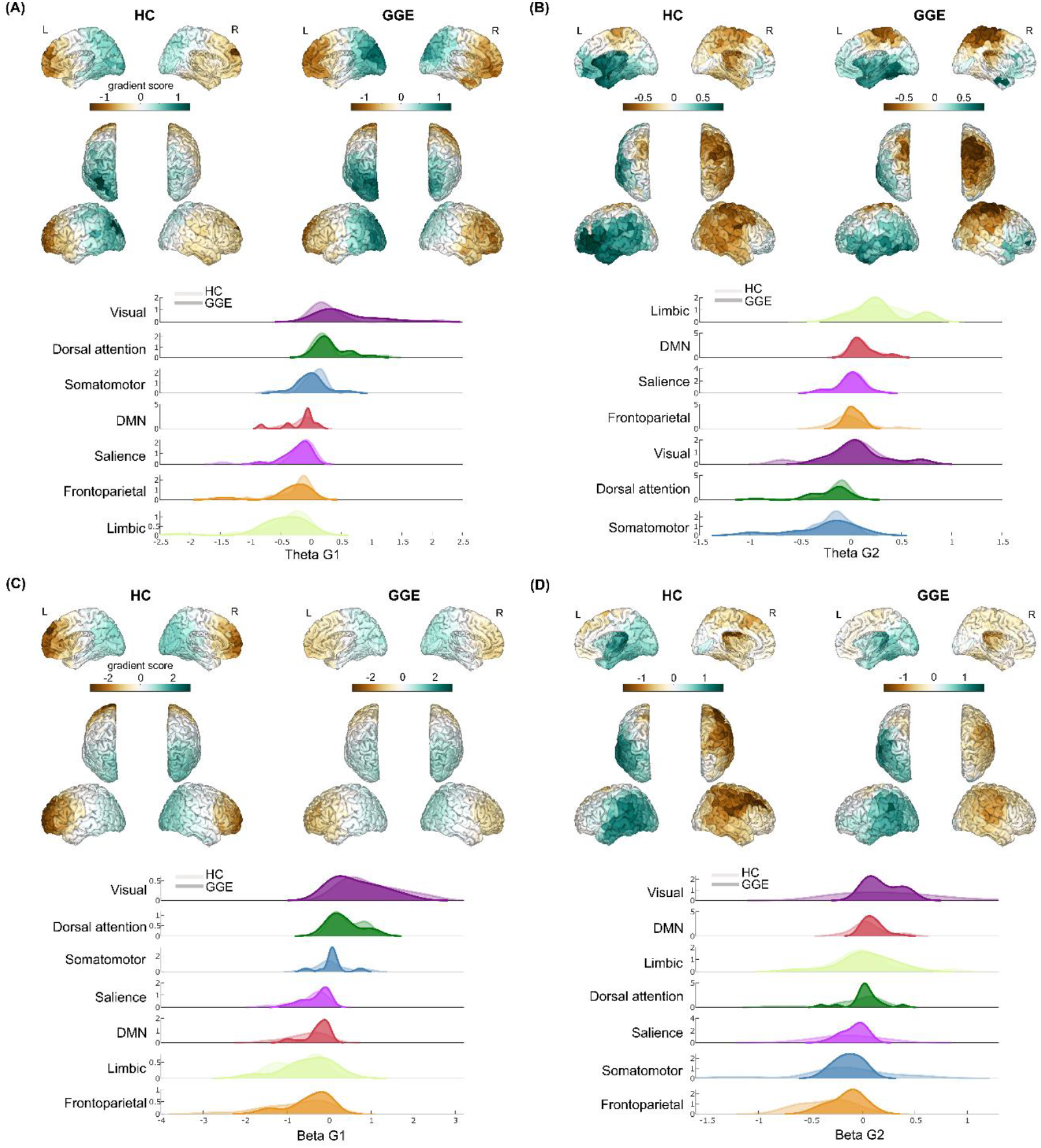
Comparison of functional network-level gradient scores from theta and beta frequency bands between the GGE and HC groups. The topographies show the group-level principal (A and C) and secondary (B and D) gradient patterns from both theta (upper panel) and beta (lower panel) frequency bands, respectively. The density plots demonstrate the distribution of the gradient scores for each functional network across all the participants within HC (in lighter color) and GGE (in darker color) groups. The color bar represents the gradient score.

Using Yeo 7 network partition, gradient scores were summarized to the sub-network level. As shown in the density plots in Figure 2, overall, the functional networks are organized with differential hierarchical patterns. In the theta band, G1 positioned the limbic and visual networks at opposite ends, with other networks in between, whilst in G2, somatomotor network (SMN) and limbic system situated at the extremes. For the beta band, G1 revealed a transition from frontoparietal network (FPN) and limbic network towards dorsal attention (DA) and visual networks, while G2 separated FPN and SMN from default mode network (DMN) and visual systems.

Statistically, we compared the groups for each network using permutation tests. As it is shown in Figure 3, in the theta band, no significant difference was found either for G1 or for G2. In the beta G1, compared to the HC, GGE group exhibited a trend for an increase (becoming more positive) in the DMN (p_uncorrected_=0.0155, p_FDR_=0.1085). Additionally, in the G2, GGE group significantly exhibited a less negative gradient score in FPN (p_uncorrected_=0.0040, p_FDR_=0.0280, effect size Cliff’s delta of 0.52), while there was no difference in the remaining networks. No difference was observed for delta or alpha band between the two groups. More details are provided in the supplementary table (Table S1). This statistical analysis confirmed significant alterations in the FPN for beta band G2 in GGE patients. Specifically, in the GGE group, while all the other networks remained unchanged, the FPN network became less negative along the second beta gradient compared to that in HC. Since the G2 situates the visual network at the positive end (note the “positive” sign simply means on one extreme direction) and the FPN on the negative end, this shift indicates that the FPN moved closer towards other networks, narrowing the global network hierarchy.

**Figure 3.**
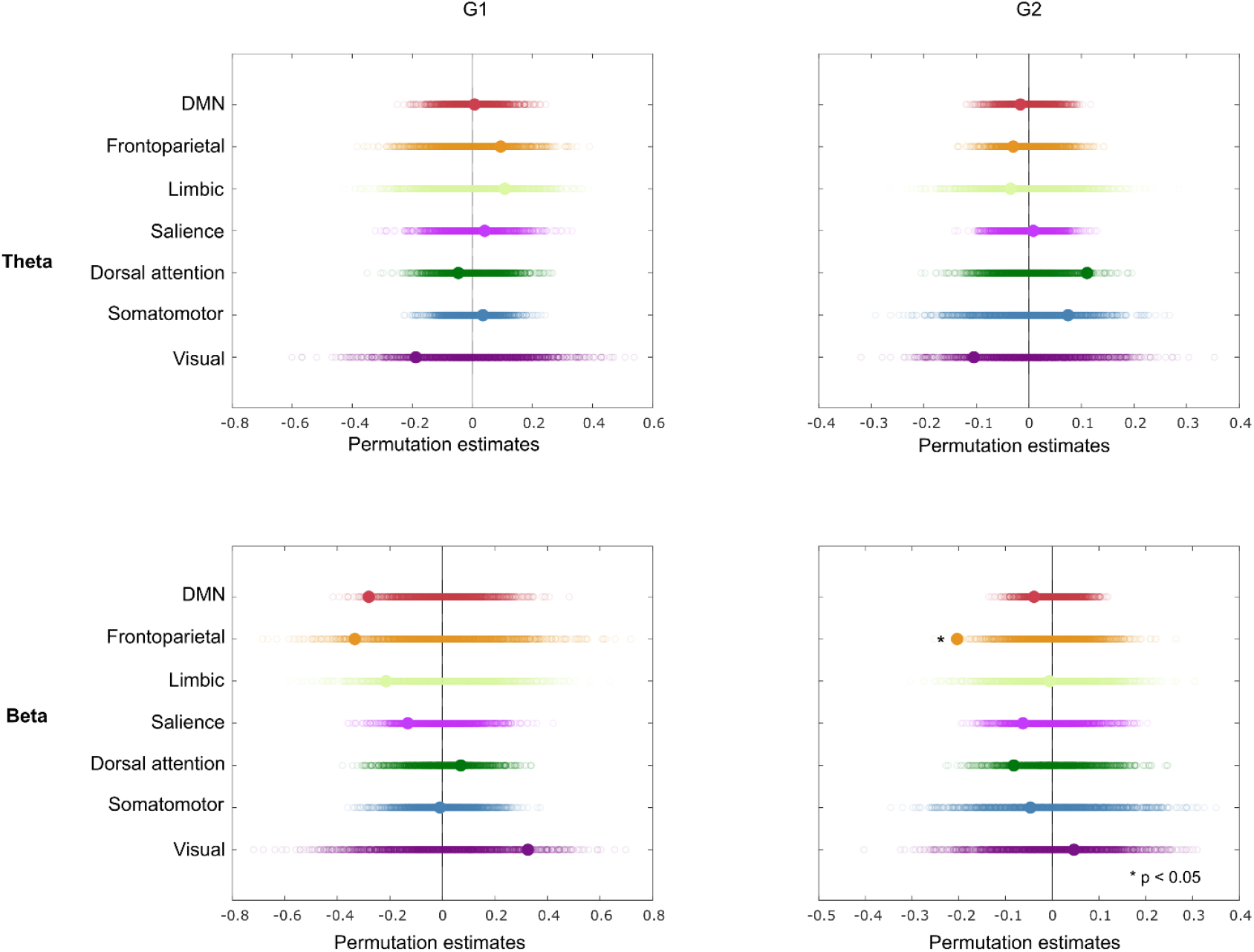
Permutation tests on the network-level gradient scores for the first two gradients from theta (upper panel) and beta (lower panel) bands. For each network, the circle represents the difference in the means between two groups (HC - GGE) of each permutation. The filled circle represents the actual difference in means among the permuted distribution. * indicates p<0.05 after FDR correction.

### 3.3 Gradient scores and clinical relevance

We investigated the relationship between the gradient scores and epilepsy duration as an index of disease severity. In the beta band, neither the principal nor the secondary gradients of all the sub-networks correlated with epilepsy duration (Table 1). In the theta band, the principal gradient showed no correlations whilst G2 significantly correlated with the epilepsy duration (age controlled) in all networks except the DMN and FPN (Spearman’s *R*=0.5093, -0.4991, -0.6108, -0.7031, 0.5278, - 0.0146, 0.1981 for visual, SMN, DA, SA, Limbic, FPN and DMN, respectively). Scatter plots for two exemplary networks are shown in Figure 4: patients with higher disease duration corresponded to more extreme gradient scores. Additionally, correlation analyses between the gradient scores and drug load showed no significant correlations (theta G2: *R*=0.0102, -0.0658, 0.0857, 0.0758, 0.0846, 0.2523, 0.1351, respectively).

**Table 1.**
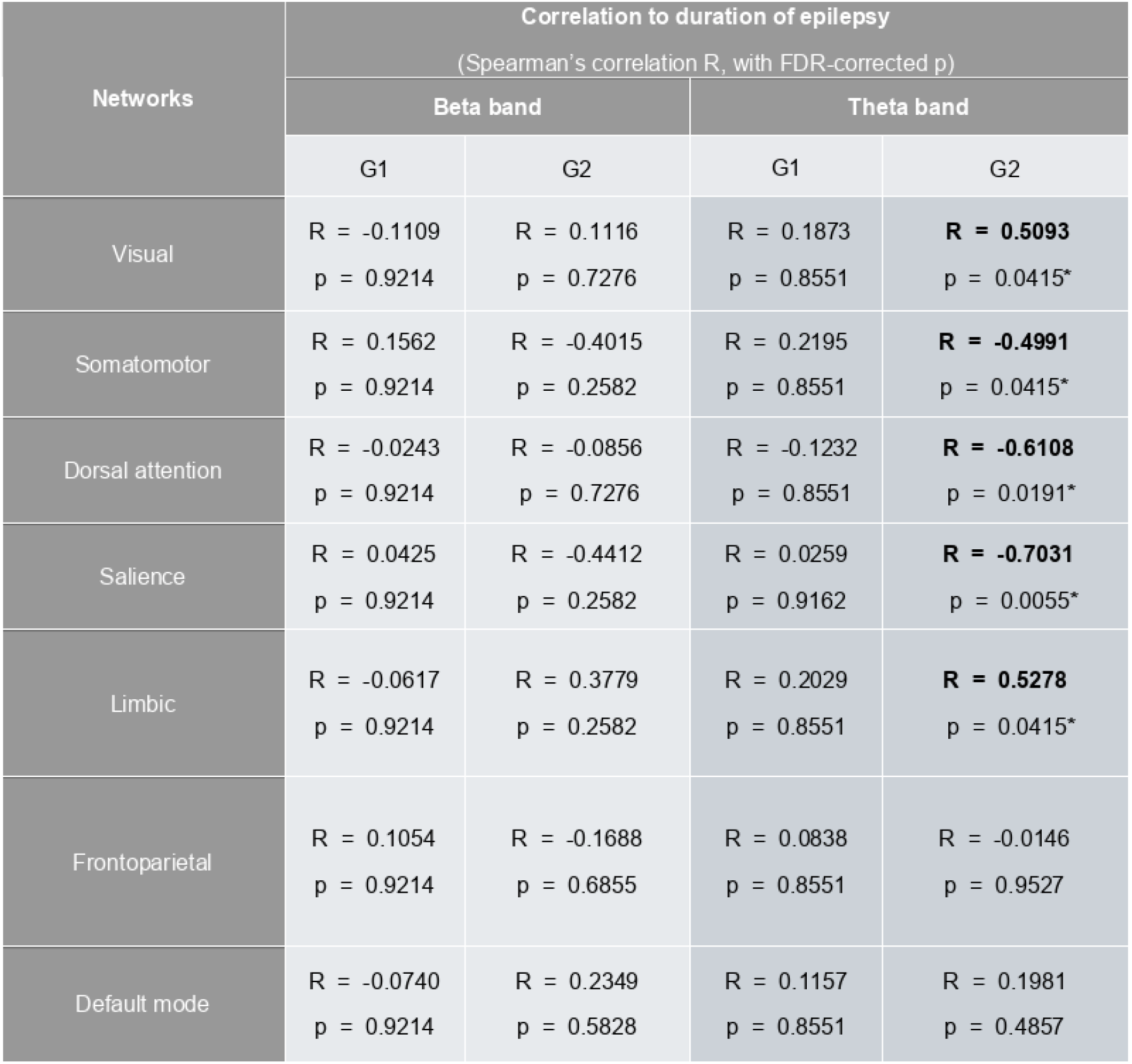
Correlations between gradient scores and duration of epilepsy

| Networks | Correlation to duration of epilepsy<br>(Spearman's correlation R, with FDR-corrected p) |  |  |  |
| --- | --- | --- | --- | --- |
|  | Beta band |  | Theta band |  |
|  | G1 | G2 | G1 | G2 |
| Visual | R = -0.1109<br>p = 0.9214 | R = 0.1116<br>p = 0.7276 | R = 0.1873<br>p = 0.8551 | **R = 0.5093**<br>p = 0.0415* |
| Somatomotor | R = 0.1562<br>p = 0.9214 | R = -0.4015<br>p = 0.2582 | R = 0.2195<br>p = 0.8551 | **R = -0.4991**<br>p = 0.0415* |
| Dorsal attention | R = -0.0243<br>p = 0.9214 | R = -0.0856<br>p = 0.7276 | R = -0.1232<br>p = 0.8551 | **R = -0.6108**<br>p = 0.0191* |
| Salience | R = 0.0425<br>p = 0.9214 | R = -0.4412<br>p = 0.2582 | R = 0.0259<br>p = 0.9162 | **R = -0.7031**<br>p = 0.0055* |
| Limbic | R = -0.0617<br>p = 0.9214 | R = 0.3779<br>p = 0.2582 | R = 0.2029<br>p = 0.8551 | **R = 0.5278**<br>p = 0.0415* |
| Frontoparietal | R = 0.1054<br>p = 0.9214 | R = -0.1688<br>p = 0.6855 | R = 0.0838<br>p = 0.8551 | R = -0.0146<br>p = 0.9527 |
| Default mode | R = -0.0740<br>p = 0.9214 | R = 0.2349<br>p = 0.5828 | R = 0.1157<br>p = 0.8551 | R = 0.1981<br>p = 0.4857 |

**Figure 4.**
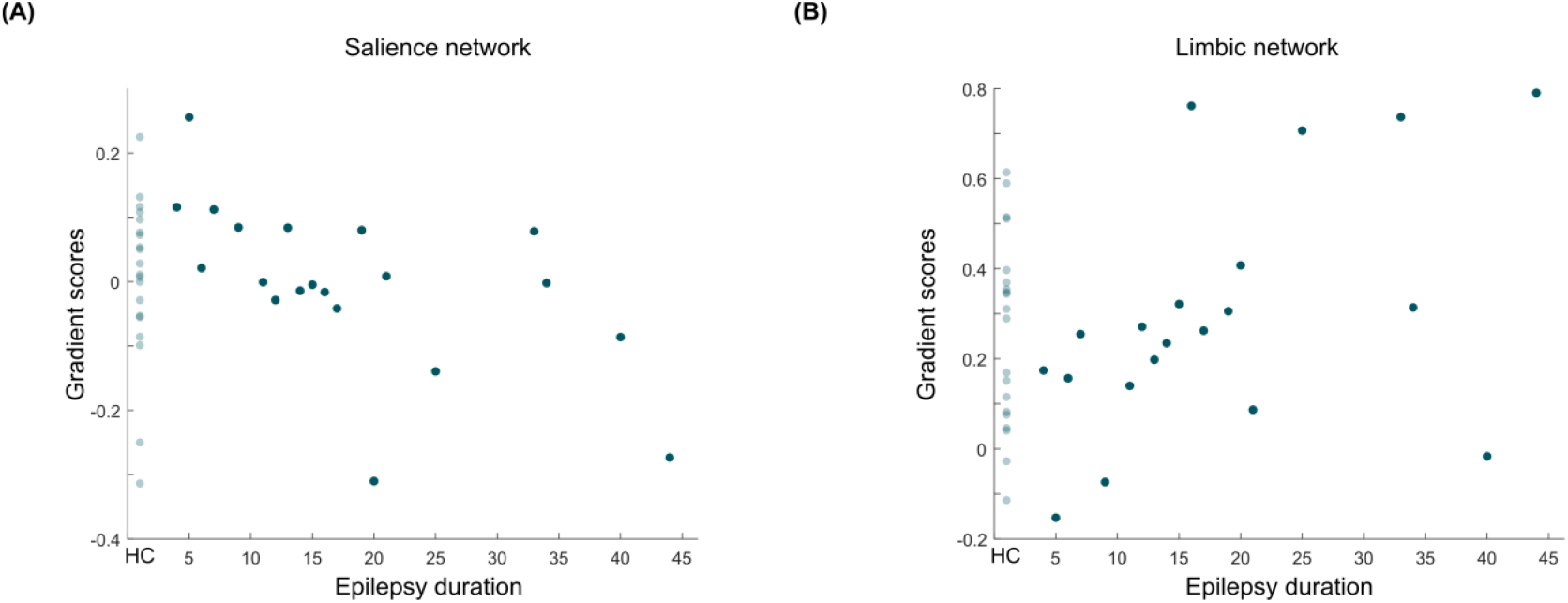
Secondary gradient scores from theta band correlated with epilepsy duration (age controlled) for all networks except DMN and FPN. Results for the salience network (A) and the limbic network (B) are shown here as examples. The light blue dots (left) represent HC samples (epilepsy duration is “0”) while darker blue dots represent the patient samples. For the SA network, the patients with longer disease duration corresponded to more negative gradient scores (*R*=-0.7031, p_FDR_=0.0055) while for the limbic network, a positive relationship was observed (*R*=0.5278, p_FDR_=0.0415).

### 3.4 Validation of main findings and control analyses

As a final step, we examined the robustness of the main findings across a wide range of thresholds (30^th^ to 90^th^ percentile). Gradient maps showed high spatial correspondence across thresholds (Figure S1). Beta-band difference effect and theta-band clinical associations (Figure S2) remained stable across thresholds, as did non-significant drug-load correlations (Figure S3). Furthermore, beta-band effect was independent of gradient component variances (Figure S4) or affinity magnitudes (Supplementary Control Analyses), confirming the reliability of the results.

## 4 Discussion

Using hdEEG, we investigated functional network gradients in GGE patients as a measure of the functional connectome hierarchy. Key findings revealed altered beta-band gradients in the FPN compared to HC. Additionally, theta-band gradient scores of multiple networks were associated with epilepsy duration. These findings remained robust against various thresholds, with control analyses confirming that the group-difference effect was independent of explained variances or the affinity magnitudes.

### 4.1 Re-positioning of the frontoparietal network along the secondary gradient axes in GGE

Our study revealed FPN gradient abnormalities in GGE, aligning with characteristic executive dysfunctions in this condition^36,37^. The FPN shifted along the secondary gradient axis (from negative to positive values) while other networks remained stable, suggesting a narrowed hierarchical network regime in GGE. This reduced inter-network distance (along the gradient axis) may facilitate pathological discharge propagation (perhaps by co-activation of FPN and epileptic networks) characterizing GGE. Notably, the effect was specific to beta band – potentially explaining executive dysfunction given its role in local processing^38^. In addition, the beta band FPN gradient alteration is aligned with our previous study, which demonstrated balanced local reorganizations between frontal and parietal regions in GGE patients^11^.

### 4.2 Gradient scores of the functional networks with its clinical relevance

Alterations in gradient scores across multiple networks correlated with duration of epilepsy while controlling for age, suggesting a progressive network hierarchy disruption as epilepsy advances. This aligns with prior fMRI findings^28^ but provides further insights regarding the network involvement of the disruption. Theta-band gradients in visual, SMN, DA, and limbic networks were correlated with clinical severity in GGE, implicating a close interaction between these regions and pathways involved in generation and maintaining of theta oscillation (e.g., the hippocampus and thalamo-cortical circuit)^39,40^, concordant with the thalamo-cortical network disorder postulated in GGE. Conversely, higher-order networks (FPN and DMN) showed no association, possibly suggesting a segregation of these networks from the other networks during the accumulation of gradient changes associated with epilepsy. This divergence might be due to increasing decoupling/dissociation of higher order functional networks from microstructural gradients present even in the healthy brain, enabling functional diversity and flexibility^13^. Future studies could investigate whether the specific re-positioning of networks occurs preceding the seizure onset. Additionally, our data showed that the drug load did not correlate with the gradient scores, ruling out treatment effects as the primary driver. The observed close association between gradients and epilepsy duration might potentially indicate its clinical significance in tracking the disease progression and suggesting therapeutic targets for non-invasive neuromodulation to restore network hierarchy and associated cognitions.

### 4.3 EEG- and fMRI-connectomes informed gradients

This study presents the first EEG-derived connectome gradient maps across frequency bands. EEG-based gradients reveal distinct patterns from fMRI-based gradients—which typically reflect the unimodal to transmodal network transition and visual-to-somatomotor separations (Figure S5)^16,19^. At the ROI level, the principal EEG-derived gradient follows a posterior-to-anterior axis, while the secondary gradient captures left-to-right hemispheric shift (supporting the hemispheric asymmetry of human cortical functional organization associated with a genetic basis)^21^. Although these EEG-based gradient maps reveal distinct patterns from what can be obtained by simple SVD decomposition on EEG data directly (instead of decomposing the connectivity patterns in the present study), they might add to the notion that analysis of resting state EEG topography may provide important information on the pathological status^41^. Network-level analysis shows the principal gradient transitioning from visual to limbic and frontoparietal networks, with the secondary gradient separating the somatomotor from limbic networks (theta band) and visual from the frontoparietal networks (beta band). Therefore, the first two EEG-derived gradient maps capture seemingly distinct distribution patterns as compared to the canonical fMRI ones (unimodal-to-transmodal and visual-to-somatomotor patterns). These differences likely arise from modality-(and possibly scale-) specific underlying brain network properties^14^. Beyond these differences, EEG-based gradients may complement the fMRI gradients, enriching the understanding of the brain spatial organization, as EEG- and fMRI-based connectomes exhibit moderate frequency-dependent cross-modal associations, as shown by studies of simultaneous EEG and fMRI recordings^42^.

To date, only few fMRI studies have examined gradient alterations in epilepsy. One study on GGE patients found reduced gradient in somatomotor areas and increased gradients in higher-order networks such as DMN and FPN, suggesting abnormal segregation between primary and higher-order networks^28^. Another recent study linked macroscale gradient differences to microcircuit dysfunction (weakened recurrent connections) highlighting the role of microcircuit dynamics in shaping epileptic network organization^43^. Similarly, the temporal lobe epilepsy (TLE) patients show expanded subcortical gradients (e.g., hippocampus and thalamus) implying a more segregated brain network^44^. Together, these findings suggest an extended gradient map in epilepsy, reflecting a dispersed hierarchical network organization.

Our data partially align with these results: while beta-band networks showed narrowed network hierarchy (greater integration), theta-band networks exhibited a tendency to be extended (DA in the theta G2 as shown in Figure 2, non-significant after FDR correction). These differences and overlaps highlight the need for replication across samples. Future simultaneous EEG-fMRI studies could clarify these patterns by examining gradient maps across both modalities.

### 4.4 Methodological considerations

Several limitations should be noted. First, this sample includes diverse syndrome types, and small subgroup sample sizes necessitated aggregation. Yet some overlapping features could be expected due to a neurobiological continuum of GGE^45^. Therefore, the observed gradient patterns may represent a common, syndrome-invariant characteristic. Nevertheless, future studies should explore the syndrome-specific gradient changes.

Secondly, while no correlation was found with the drug load, antiepileptic drug effects cannot be entirely excluded. The difference effect in the beta band may reflect a complex disease-drug interaction. Future investigation, if possible, of drug-naive patients would help clarify this. Finally, these findings may be specific to the current dataset, and analyses incorporating patients with a differential disease severity spectrum might yield different theta-band gradient alterations or changes in other frequency bands.

## 5 Conclusion

Our study presents the first EEG-based evidence of GGE-related gradient patterns, with large effect size supporting potential clinical diagnostic value. We found frequency-dependent gradient signatures of narrowed network hierarchy between FPN and other networks in beta band, reflecting greater integration, possibly contributing to the pathological discharge spread and the executive dysfunction.

Theta-band gradients from multiple networks closely correlated with epilepsy duration, indicating progressive network hierarchy disruption and involvement of distributed theta-relevant networks.

## Data Availability

The data supporting the findings of this study are available upon reasonable request from the corresponding author. The data are not publicly available as they contain information which could comprise the data privacy of the participants. Derived data for reproducing the key findings are awailable here: https://figshare.com/s/0b2fd05de5ab070fe796.

## Acknowledgements

This work was supported by Swiss National Science Foundation (SNSF) grants 209120, 209470, 192749, 10004348.

## Author contributions

**Juanli Zhang**: Conceptualization; data curation; methodology; formal analysis; writing–original draft, review & editing. **Isotta Rigoni**: Data acquisition; writing–review and editing. **Dimitri Van De Ville**: writing–review & editing. **Serge Vulliémoz**: Conceptualization; supervision; funding acquisition; writing–review & editing. **Nicolas Roehri**: Conceptualization; data acquisition; methodology; supervision; funding acquisition; writing–review & editing.

