## Supplemental Material for "Disruption of functional gradients in genetic generalized epilepsy and its clinical relevance: evidence from high-density EEG"

### Supplementary material

#### 1. Validation of the main findings: across a family of thresholds

To validate the robustness of the main results (80th percentile retaining the top 20% of the connections was used), we examined the resulted gradient patterns when a family of different thresholding values were used. The thresholds varied from 30th to 90th percentile with a step of 10th (90th percentile corresponds to retaining the top 10% of the connections).

Across all the thresholds, the group-level principal and secondary gradient scores from both bands exhibit high similarities with the main results (see Figure S1). The gradient patterns showed high spatial correspondence with the patterns shown in the main results (all *R*>0.89). This high degree of similarity indicates the strong robustness of the results across different processing parameters.

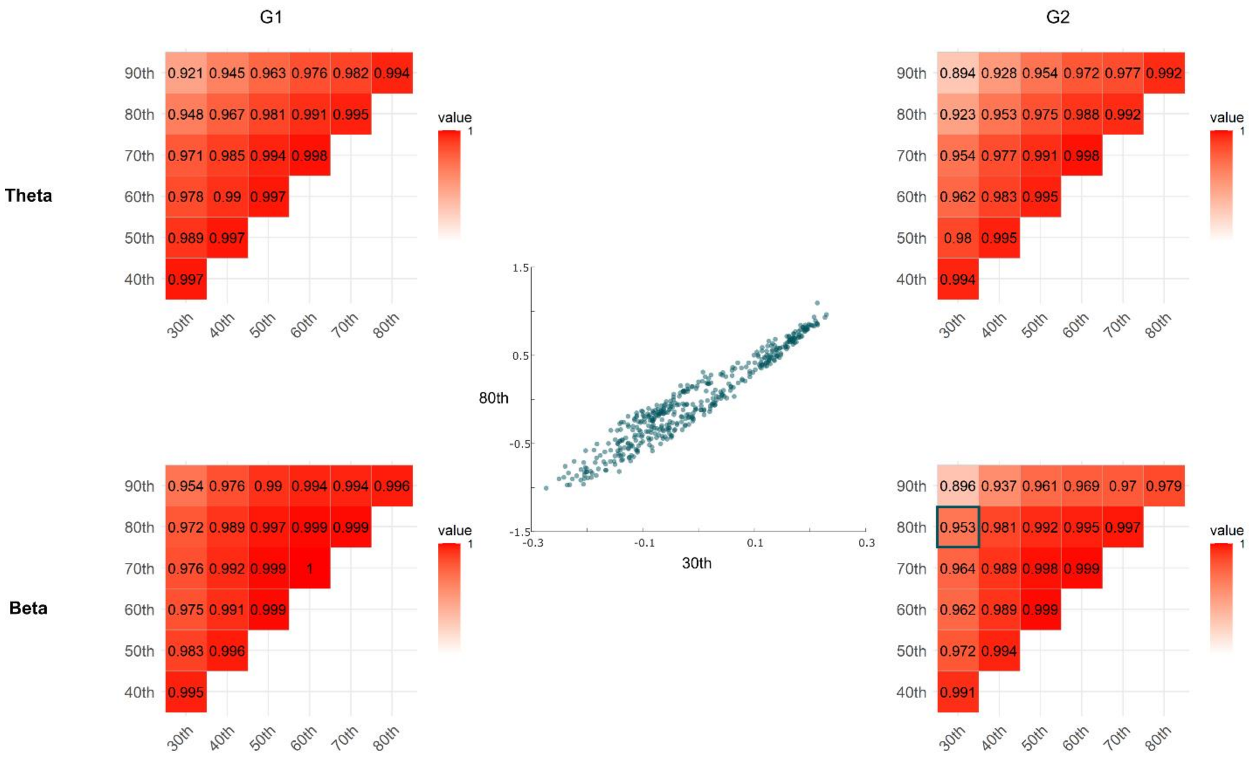


Figure S1. Spatial correspondence between the gradient patterns across a family of thresholds and the counterparts in the main result (thresholding at 80th percentile) for theta band (upper panel) and beta band (lower panel). The scatter plot in the middle shows the correspondence of the secondary beta gradient scores from the 30th- versus 80th- percentile thresholded functional connectomes.

Furthermore, we tested whether the significant alteration of the gradients from beta band as well as the significant relationships between gradients from theta band with epilepsy duration can be replicated through applying the different thresholds to the functional connectome. As it is shown in the panel A of Figure S2, across all the thresholding percentiles, in the beta band the gradient score of FPN in GGE group significantly differed from those of the HC group. As for the theta band, with the varying thresholding parameters, the secondary gradient scores from multiple networks consistently showed significant correlations with the epilepsy duration while accounting for the age (see panel B of Figure S2). The correlations from networks of SMN and DA showed consistency across all the thresholds while the visual, salience and limbic networks across most of the thresholding settings. These results further confirmed that the main findings are robust against the processing settings.

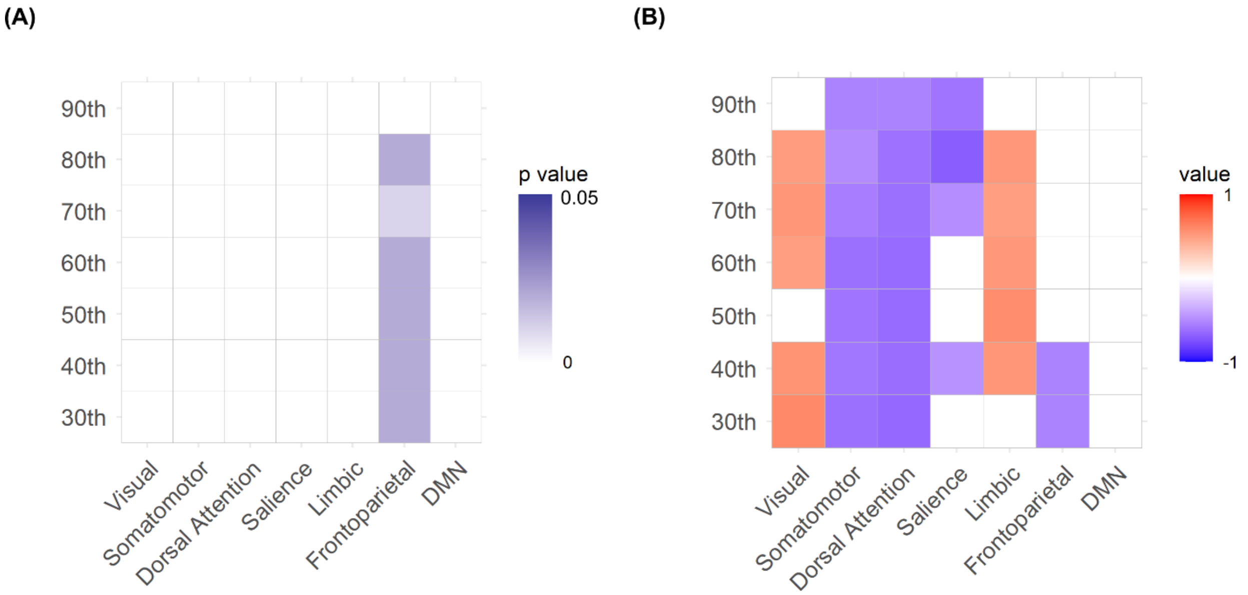


Figure S2. Stability of the significant effects across a family of thresholds (30th to 90th with a step of 10th) retaining from top 70% to 10% of the connections. (A) The p-value map (after FDR correction) for the comparisons between two groups in the secondary gradient from beta frequency band: across all the thresholds except for the 90th percentile, gradient scores of FPN were significantly different in GGE patients as compared to the HC. (B) Across all the thresholds, gradient scores of SMN, DA and salience network from G2 of theta band negatively correlated with epilepsy duration (while controlling for age) whilst gradient scores of the visual and limbic systems exhibited positive correlations with epilepsy duration across most of the thresholding parameters. The non-significant ones (p>0.05 after FDR correction) are set to zero illustrated by while color.

In addition, we sought to test whether non-significant correlations between the gradient scores and drug load were consistent across all the thresholds. As it is shown in the Figure S3, indeed, across all the thresholding percentiles, there was no significant correlation for either theta or beta band (p values were FDR-corrected).

(A)

(B)

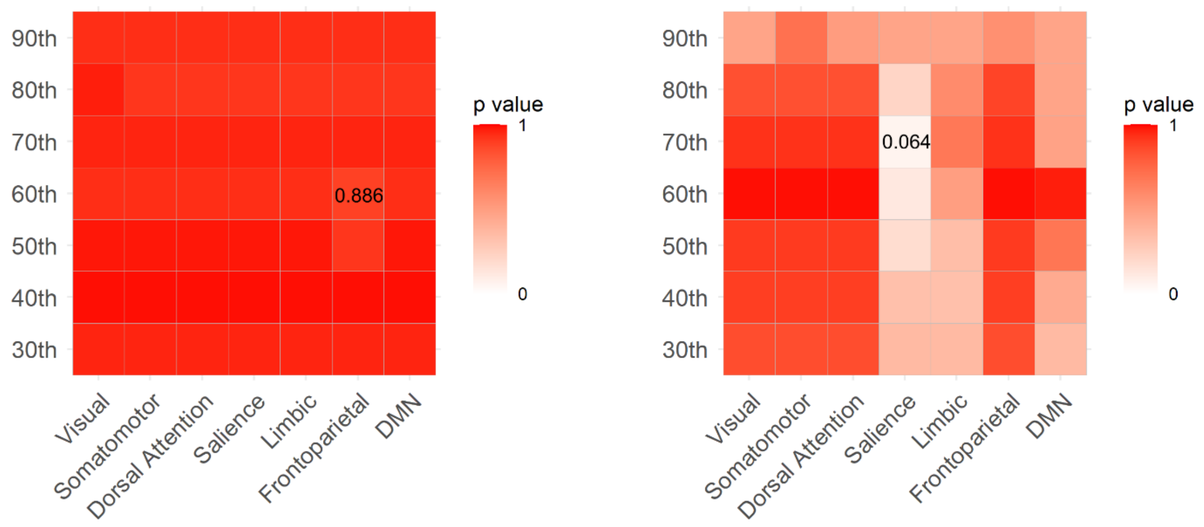


Figure S3. Stability of the non-significant correlations between the gradient scores with the drug load across a family of thresholds from 30th to 90th percentile (retaining from top 70% to 10% of the connections) for (A) theta band and (B) the beta band. All p-values were corrected using FDR. The highlighted values were the smallest values within each p-map: 0.886 for A and 0.064 for B.

### 2. Control analyses

Note, the significant alteration in gradient scores in GGE group is not driven by the connectivity strength per se as “cosine similarity” estimates the cosine of the angle between vectors thus being robust to magnitude. Yet, we performed two additional analyses to ensure the gradient changes observed between the two groups are not accounted for by other confounding factors such as the amount of the variances explained by the gradients or magnitude of the affinity metric.

As it is shown in Figure S4, the scaled eigenvalues (lambdas) of the first 20 gradient components were plotted for both groups and frequency bands. It appears a reasonable choice to keep the first two gradients as they provide a good trade-off between keeping few components while retaining a large amount of variance. Overall, the amount of explained variance (indicated by the normalized eigenvalues) between the two groups were similar across all the tested gradient components. Statistical tests revealed that there was no difference between two groups for both bands (p=0.6736 and p=0.2710 for G1 and G2, respectively, from theta band; p=0.6978 and p=0.8244 for G1 and G2, respectively, from the beta band, all uncorrected). This analysis suggested that the significant difference in gradient scores from the beta band between the two groups is unlikely to be driven by the potential bias from the lambdas.

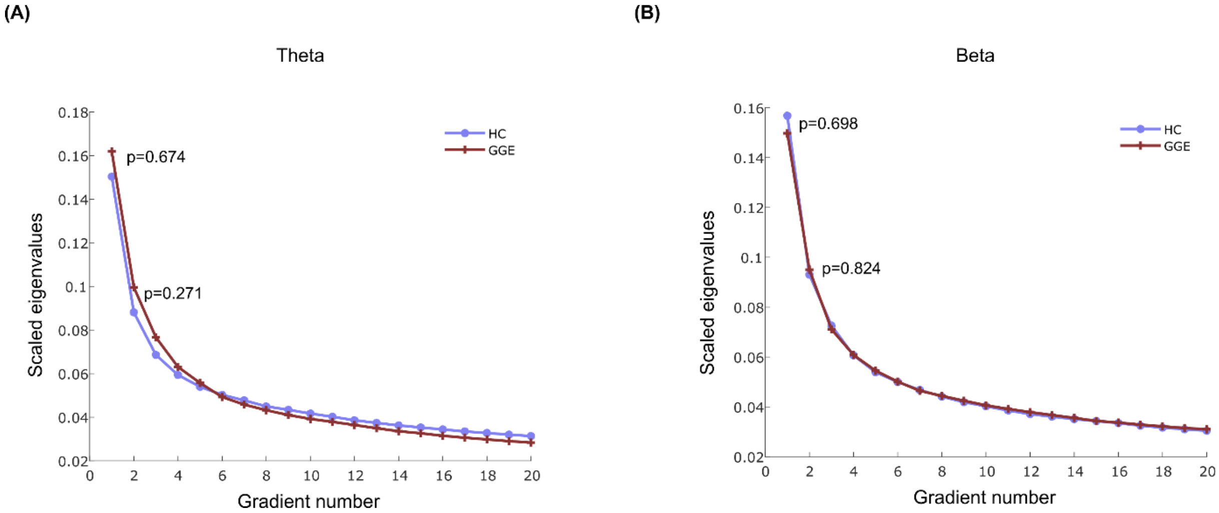


Figure S4. The distribution of explained variance by the first 20 gradients for both groups. The amounts of variances explained by gradients (G1 and G2) from both frequency bands were not significantly different between two groups.

Additionally, we summed up the values across all the ROIs within the affinity matrix for each participant and compared the overall affinity magnitude between the two groups. Statistical tests revealed that there was no significant difference in the affinity metrics between the two groups either for theta or beta frequency band (p=0.7907 and p=0.3138, respectively). This analysis result demonstrated that the change of gradient scores in GGE group in comparison with the HC is not accounted for by a possible bias from the affinity metric, which is an intermediate output of the gradient mapping.

### 3. Additional material

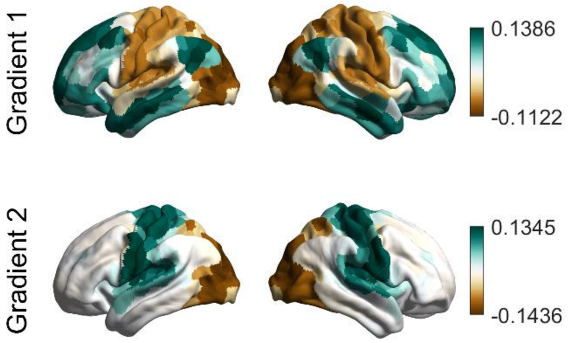


Figure S5. Typical fMRI-connectome derived gradients using BrainSpace toolbox.

Table S1. Comparison of gradient scores G1 and G2 between two groups

| Networks | Comparisons of the gradient scores (All p values are after FDR correction) | | | |
|---|---|---|---|---|
| | Beta band | | Theta band | |
| | G1 | G2 | G1 | G2 |
| Visual | p = 0.2555 | p = 0.7805 | p = 0.7169 | p = 0.7752 |
| Somatomotor | p = 0.9300 | p = 0.7805 | p = 0.7169 | p = 0.7752 |
| Dorsal attention | p = 0.6335 | p = 0.6116 | p = 0.7169 | p = 0.3115 |
| Salience | p = 0.3150 | p = 0.6116 | p = 0.7169 | p = 0.8265 |
| Limbic | p = 0.3150 | p = 0.9500 | p = 0.7169 | p = 0.7752 |
| Frontoparietal | p = 0.2555 | ***p = 0.0280** | p = 0.7169 | p = 0.7752 |
| Default mode | p = 0.1085 | p = 0.6116 | p = 0.9135 | p = 0.7752 |
